# Prediction of in-hospital mortality at a 72-hour intensive care landmark in patients with sepsis and pre-existing hypertension: development and external validation

**DOI:** 10.64898/2026.10.02.26364566

**Authors:** Yufeng Chen¹, Yuzhen Cai¹, Hongyang Bian¹, Mingyan Fu¹

**Author notes:** Corresponding author: Mingyan Fu.

## Abstract

**Background:** In patients with sepsis and pre-existing hypertension, admission findings may not adequately describe the subsequent course of circulatory support and organ dysfunction. We developed and externally validated a model to reassess in-hospital mortality risk after the first 72 hours of intensive care.

**Methods:** This retrospective study used MIMIC-IV for development and eICU-CRD for external validation. Patients had received vasoactive agents and remained in the ICU for at least 72 hours. Static variables were combined with features from six consecutive 12-hour windows. Imputation, feature selection, model fitting, and calibration were performed within training folds during five-fold cross-validation. The final XGBoost model was applied externally without refitting. Evaluation included discrimination, calibration, decision curves, and risk stratification, with an exploratory comparison of static-only and static-plus-dynamic feature sets.

**Results:** The development and external cohorts included 1019 and 1747 patients, with 338 (33.2%) and 516 (29.5%) deaths, respectively. Internal area under the receiver operating characteristic curve (AUC) was 0.752 (95% CI 0.720–0.784). External AUC was 0.713 (95% CI 0.687–0.739), average precision was 0.536, the Brier score was 0.185, and the calibration slope was 0.744. Mortality increased across external risk tertiles from 13.7% to 47.3%. In an exploratory feature-set comparison, the static-plus-dynamic feature set showed a 0.071 higher internal AUC than the static feature set (95% CI 0.042–0.109).

**Conclusions:** Information accumulated during the first 72 hours supported subsequent mortality risk stratification in this selected population. The model retained moderate discrimination externally, supporting further evaluation as an adjunct to clinical reassessment. Local calibration and the clinical consequences of acting on model predictions require evaluation before routine use.

## Introduction

Sepsis remains an important cause of death despite advances in recognition and supportive care [1–3]. Among patients receiving vasopressors, prognosis reflects the course of organ dysfunction and treatment requirements as well as the severity of hypotension [4–5]. Pre-existing hypertension adds a relevant clinical context because chronic vascular changes and altered autoregulation may influence organ perfusion during acute circulatory illness [6–8]. Blood-pressure trials have not established a consistent survival advantage from a higher target [9–12]. In the hypertension subgroup of SEPSISPAM, however, the renal findings highlighted the need to consider organ responses alongside arterial pressure [9]. These observations provide a rationale for studying prognosis in patients with this combination of chronic disease and acute illness.

For patients who remain in the ICU after initial resuscitation, the practical question is how to reassess risk as the clinical course unfolds. By 72 hours, repeated measurements of urine output, blood pressure, lactate, and vasoactive-agent requirements describe aspects of persistent illness or recovery that are not captured by admission measurements alone. Previous studies have used admission variables or evolving physiological data to predict mortality in sepsis and critical illness [13–15]. A 72-hour landmark offers a defined opportunity to examine the value of accumulated information for subsequent prognosis, distinct from early sepsis recognition [16]. In patients with pre-existing hypertension, assessing pressure measurements together with organ responses and support requirements may be particularly relevant to this reassessment.

We therefore examined whether static clinical variables combined with information from the first 72 hours could stratify subsequent in-hospital mortality risk among patients with sepsis and pre-existing hypertension who had received vasoactive agents. We developed the model in MIMIC-IV and evaluated it independently in eICU-CRD. Discrimination, calibration, decision curves, and risk stratification were assessed, and an exploratory feature comparison examined the additional predictive information associated with the combined dynamic feature set.

## Materials and methods

### Study design and participants

We conducted a retrospective study using MIMIC-IV version 3.1 for development and eICU-CRD version 2.0 for external validation [17–18]. The collection and sharing of MIMIC-IV data were reviewed by the Institutional Review Board at Beth Israel Deaconess Medical Center, which approved the data-sharing initiative and granted a waiver of informed consent. eICU-CRD contains de-identified data distributed through PhysioNet under credentialed access, with access requiring human-subjects research training and a signed data use agreement. Investigators completed the required PhysioNet training and credentialing and signed the applicable data use agreement (Record ID 50141015). The data were accessed for research purposes on 15 November 2025. This secondary analysis involved no patient contact and no access to identifiable information.

Patients with sepsis and pre-existing hypertension were identified separately in the MIMIC-IV and eICU-CRD databases using prespecified diagnosis codes or standardized diagnostic records. In MIMIC-IV, sepsis was identified using ICD-9 codes 995.91, 995.92, and 785.52, and pre-existing hypertension was identified using ICD-9 codes 401.0, 401.1, 401.9, 402.xx, 403.xx, and 404.xx. The eICU-CRD cohort was identified according to the available standardized diagnostic records using the same clinical framework. To improve cohort specificity and favor inclusion of patients with organ dysfunction evident early in the ICU course, patients were additionally required to have a Sequential Organ Failure Assessment (SOFA) score of at least 2 within the first 24 hours after ICU admission [19–20]. We included the first ICU stay within a hospital admission, with an ICU stay of at least 72 hours and vasoactive-agent exposure during that window. Patients younger than 18 or older than 89 years and records with more than 20% missing candidate-variable data were excluded. Each cohort contained one admission record per patient, without repeated admissions. The prediction landmark was 72 hours after ICU admission, and the outcome was subsequent in-hospital death.

Predictors included demographic characteristics, comorbidities, initial vital signs and laboratory measurements, severity scores, and treatment-related variables. Repeated measurements during the first 72 hours were assigned to six consecutive 12-hour windows. We derived level summaries (first and last observed values, mean, minimum, and maximum), temporal changes (early-to-late difference and linear slope), variability (coefficient of variation and adjacent-window changes), and clinically defined burden or treatment-response measures when applicable. Unobserved windows remained missing until imputation. Observation windows, variable definitions, and model-input units were harmonized across databases; S1 Text gives the dynamic-feature definitions and formulas.

### Missing data and feature selection

Missingness was assessed before imputation. A chained-equation procedure was fitted separately within each training partition of five-fold cross-validation and applied unchanged to its held-out partition. The 20% exclusion criterion applied to admission records, not individual predictors. No predictor was excluded solely on the basis of its variable-level missingness.

Feature selection was repeated within each training fold. Boruta compared candidate predictors with randomly permuted shadow features, after which the least absolute shrinkage and selection operator (LASSO) reduced the correlated feature set. Each fold-specific model generated predictions for its held-out patients. Selection stability was assessed using feature counts, selection frequencies, and pairwise Jaccard similarity. A single predictor set was required for fitting the final model to the full development cohort. We therefore used the union of features selected in at least one training fold, while characterizing selection stability separately through selection frequencies and pairwise Jaccard similarity. The resulting predictor set was used for full-cohort fitting and external evaluation.

### Model development

We compared extreme gradient boosting (XGBoost), LightGBM, random forest, support vector machine, logistic regression, a back-propagation neural network, and k-nearest neighbors. Sigmoid calibration was also evaluated for XGBoost to adjust predicted probabilities. Imputation, feature selection, model fitting, and calibration were performed within the training partitions of five-fold cross-validation, producing one held-out prediction per patient. SAPS II (Simplified Acute Physiology Score II) [21] and the first SOFA score provided admission-window clinical references. Implementation settings and software dependencies will accompany the publicly released code.

### Model interpretation

We used Shapley additive explanations (SHAP) to describe the model fitted to the full development cohort [22]. Mean absolute attribution was summarized for individual predictors and for baseline or static, 72-hour level or summary, and trajectory or variability features. Each category was expressed as a proportion of total mean absolute attribution.

### Internal performance evaluation

Discrimination was evaluated using the area under the receiver operating characteristic curve (ROC AUC) and average precision; overall prediction error was assessed using the Brier score. Calibration intercepts and slopes were estimated by regularized logistic regression of the outcome on the logit of predicted risk. Unless otherwise specified, 95% confidence intervals used 2000 patient-level bootstrap resamples of fixed predictions. Exploratory sensitivity, specificity, predictive values, F1 score, and net benefit were calculated from pooled out-of-fold predictions at three reference thresholds. Decision curves compared the model with treat-all and treat-none strategies [23–24]. Mortality across tertiles of predicted risk was summarized descriptively.

### External validation

The final sigmoid-calibrated XGBoost model was fitted to the full MIMIC-IV cohort using the union of selected predictors, then locked and applied to eICU-CRD without refitting or recalibration. Discrimination, prediction error, calibration, decision-curve analysis, and risk stratification were assessed in the external cohort. External decision curves covered threshold probabilities of 0.05–0.30, and risk tertiles were defined within the external predicted-risk distribution.

### Exploratory feature comparison

We compared static-only and static-plus-dynamic feature sets within the development cohort using identical five-fold patient splits. Imputation, Boruta–LASSO selection, XGBoost fitting, and sigmoid calibration were repeated independently within each training fold of each arm. We compared pooled out-of-fold AUC, average precision, and Brier score. Confidence intervals for the paired comparison used 2000 patient-level bootstrap resamples of fixed predictions. This two-arm comparison was selected after initial exploration of candidate feature sets and was considered post hoc.

### Statistical analysis

Categorical variables are reported as counts and percentages and were compared using the chi-square test or Fisher’s exact test. Continuous variables are reported as medians and interquartile ranges and were compared using the Wilcoxon rank-sum test. MIMIC-IV variables were extracted using pgAdmin 4 version 8.13 and Navicat Premium version 17.0.8. Variables from eICU-CRD were extracted and harmonized according to the development-cohort definitions. Python version 3.10 was used for machine-learning modeling, external validation, decision curve analysis, risk stratification, and SHAP analyses. Statistical analyses were performed in SPSS version 29.0.2.0. No a priori sample-size calculation was performed because all eligible patients meeting the prespecified criteria in each database were included. A two-sided P < 0.05 was considered statistically significant.

## Results

### Cohort characteristics

The MIMIC-IV development cohort included 1019 patients, of whom 338 (33.2%) died after the landmark. Cohort selection is shown in Fig 1. The eICU-CRD external validation cohort included 1747 patients, with 516 deaths (29.5%). In the development cohort, nonsurvivors had greater comorbidity burden and organ dysfunction, lower blood pressure and urine output, and higher lactate and norepinephrine doses (Table 1). Median cumulative 72-hour urine output was 2301.5 mL [792.5, 4606.8] in nonsurvivors and 4528.5 mL [2855.3, 6778.8] in survivors. External cohort characteristics are presented in S3 Table.

**Fig 1.**
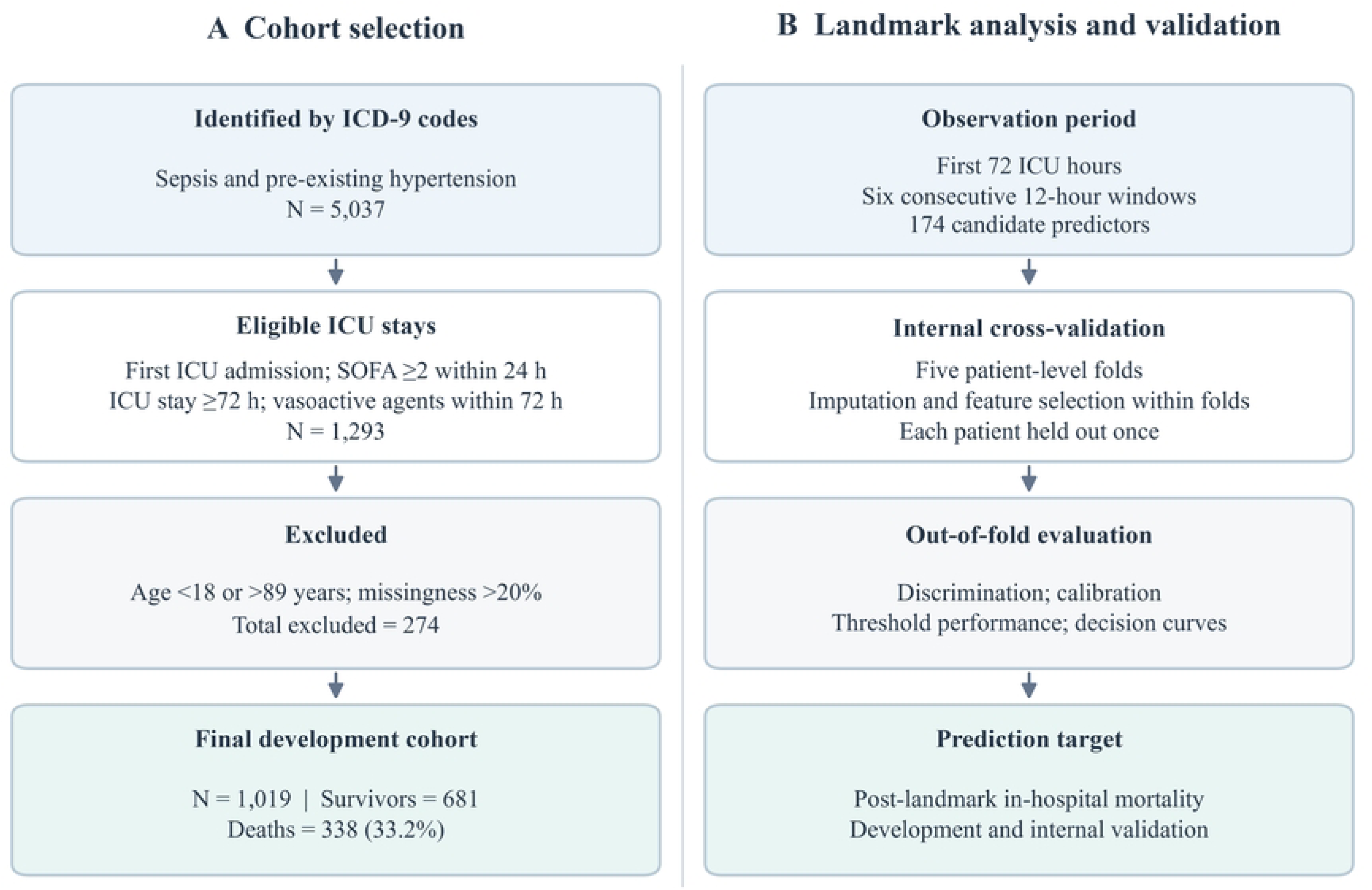
Flowchart of cohort selection and the 72-hour landmark analysis. (A) Selection of the MIMIC-IV development cohort. (B) The 72-hour observation window and five-fold internal validation workflow. The flowchart covers model development and internal validation. ICD-9, International Classification of Diseases, Ninth Revision; ICU, intensive care unit; SOFA, Sequential Organ Failure Assessment; OOF, out-of-fold.

**Table 1.** Admission characteristics and clinical summaries from the first 72 hours in the MIMIC-IV development cohort, stratified by subsequent in-hospital death.

| Characteristic | Overall (n = 1019) | Survivors (n = 681) | Deaths (n = 338) | P value |
| --- | --- | --- | --- | --- |
| Age, years | 70.39 [61.27, 79.75] | 69.40 [59.90, 78.73] | 73.18 [63.12, 81.14] | <0.001 |
| Male sex | 569 (55.8) | 392 (57.6) | 177 (52.4) | 0.116 |
| Body mass index, kg/m<sup>2</sup> | 29.14 [24.91, 34.89] | 29.36 [25.25, 35.30] | 28.46 [24.39, 34.17] | 0.114 |
| Chronic kidney disease | 397 (39.0) | 254 (37.3) | 143 (42.3) | 0.123 |
| Malignancy | 137 (13.4) | 67 (9.8) | 70 (20.7) | <0.001 |
| Acute kidney injury | 929 (91.2) | 602 (88.4) | 327 (96.7) | <0.001 |
| KDIGO stage | 2.00 [2.00, 3.00] | 2.00 [2.00, 3.00] | 3.00 [2.00, 3.00] | <0.001 |
| Invasive mechanical ventilation | 790 (77.5) | 514 (75.5) | 276 (81.7) | 0.026 |
| Continuous renal replacement therapy | 125 (12.3) | 64 (9.4) | 61 (18.0) | <0.001 |

| **Characteristic** | **Overall (n = 1019)** | **Survivors (n = 681)** | **Deaths (n = 338)** | **P value** |
| --- | --- | --- | --- | --- |
| Multiple vasoactive agents | 363 (35.6) | 210 (30.8) | 153 (45.3) | <0.001 |
| Charlson Comorbidity Index | 6.00 [4.00, 8.00] | 6.00 [4.00, 8.00] | 7.00 [5.00, 9.00] | <0.001 |
| Platelet count, $\times 10^9/L$ | 192.00 [126.00, 291.00] | 201.50 [139.00, 299.00] | 172.00 [107.00, 275.00] | <0.001 |
| Mean systolic blood pressure, mmHg | 109.11 [103.50, 115.69] | 110.25 [104.95, 116.83] | 105.86 [100.61, 111.84] | <0.001 |
| Mean arterial pressure, mmHg | 71.97 [67.82, 76.75] | 73.20 [68.83, 78.15] | 70.31 [66.23, 74.48] | <0.001 |
| Mean lactate, mmol/L | 1.81 [1.30, 2.74] | 1.64 [1.18, 2.32] | 2.26 [1.55, 3.44] | <0.001 |
| Mean base excess, mmol/L | -2.88 [-5.83, 0.00] | -2.50 [-5.42, 0.00] | -3.50 [-6.83, -0.85] | <0.001 |
| Mean 12-hour urine output, mL | 652.67 [312.29, 1011.75] | 757.50 [479.71, 1132.50] | 387.08 [135.23, 773.50] | <0.001 |
| Cumulative 72-hour urine output, mL | 3914.00 [1848.00, 6070.50] | 4528.50 [2855.25, 6778.75] | 2301.50 [792.50, 4606.75] | <0.001 |
| Mean norepinephrine dose, $\mu g/kg/min$ | 0.04 [0.01, 0.10] | 0.03 [0.01, 0.08] | 0.06 [0.02, 0.13] | <0.001 |
| Mean number of active vasoactive agents | 0.83 [0.50, 1.17] | 0.67 [0.33, 1.00] | 1.00 [0.67, 1.33] | <0.001 |

The development analysis considered 174 candidate predictors. Their missingness before imputation is summarized in S2 Fig.

Continuous variables are shown as median [first quartile, third quartile] and compared using the Wilcoxon rank-sum test; categorical variables are shown as n (%) and compared using the chi-square test or Fisher’s exact test. All P values are two-sided. KDIGO, Kidney Disease: Improving Global Outcomes.

### Selected predictors

Boruta–LASSO retained 23, 21, 18, 29, and 26 predictors across the five training folds. Seven were selected in every fold and 11 in only one; mean pairwise Jaccard similarity was 0.410 (Fig 2). The consistently selected predictors were age, Charlson Comorbidity Index, minimum and last systolic blood pressure, last norepinephrine dose, minimum urine output, and log-transformed late-to-early urine-output ratio. The union contained 45 predictors and was used for full-cohort fitting, SHAP analysis, and external validation.

**Fig 2.**
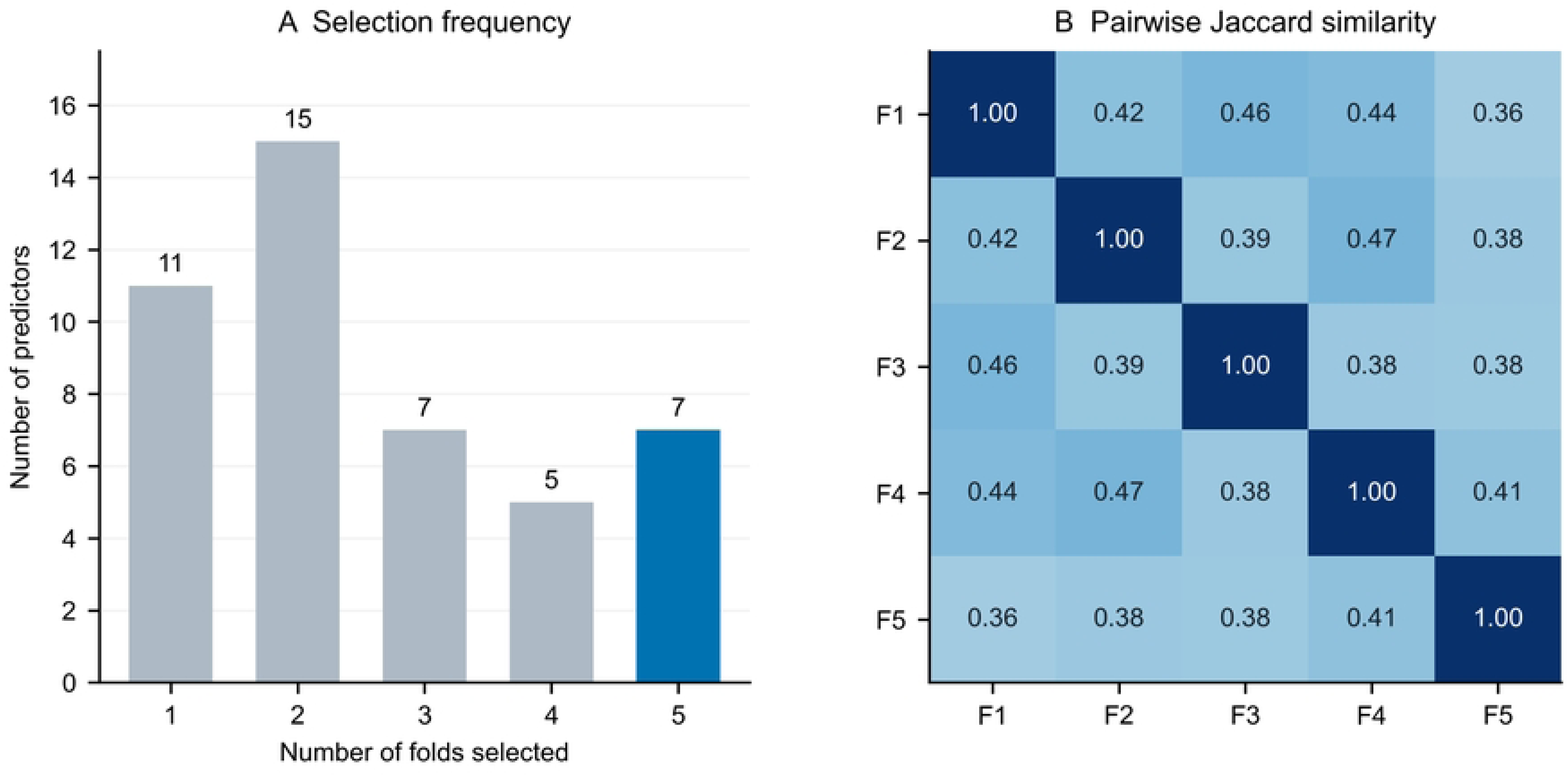
Stability of feature selection across training folds. (A) Selection frequencies of the 45 predictors across one to five training folds; seven were retained in all folds (blue). (B) Pairwise Jaccard similarity between fold-specific feature sets (F1–F5). Diagonal cells indicate self-comparison, and off-diagonal cells indicate overlap between folds. Boruta–LASSO selection was repeated independently within each training fold. LASSO, least absolute shrinkage and selection operator.

### Internal model performance

Sigmoid-calibrated XGBoost yielded an out-of-fold AUC of 0.752 (95% CI 0.720–0.784), average precision of 0.610, and Brier score of 0.182 (Fig 3; Table 2). The event proportion was 0.332. Uncalibrated XGBoost had an AUC of 0.751, Brier score of 0.186, and calibration slope of 0.670; the calibrated slope was 0.966. Paired AUC differences relative to uncalibrated XGBoost and random forest were 0.001 (95% CI −0.006 to 0.008) and 0.011 (−0.010 to 0.030), respectively. AUCs for SAPS II and first SOFA were 0.651 and 0.615. Exploratory operating characteristics across three reference thresholds are summarized in Table 3A and illustrate the expected trade-off between sensitivity and specificity. Observed mortality increased across internal low-, intermediate-, and high-risk tertiles from 14.1% to 29.2% and 56.2%, respectively (Table 3B).

**Fig 3.**
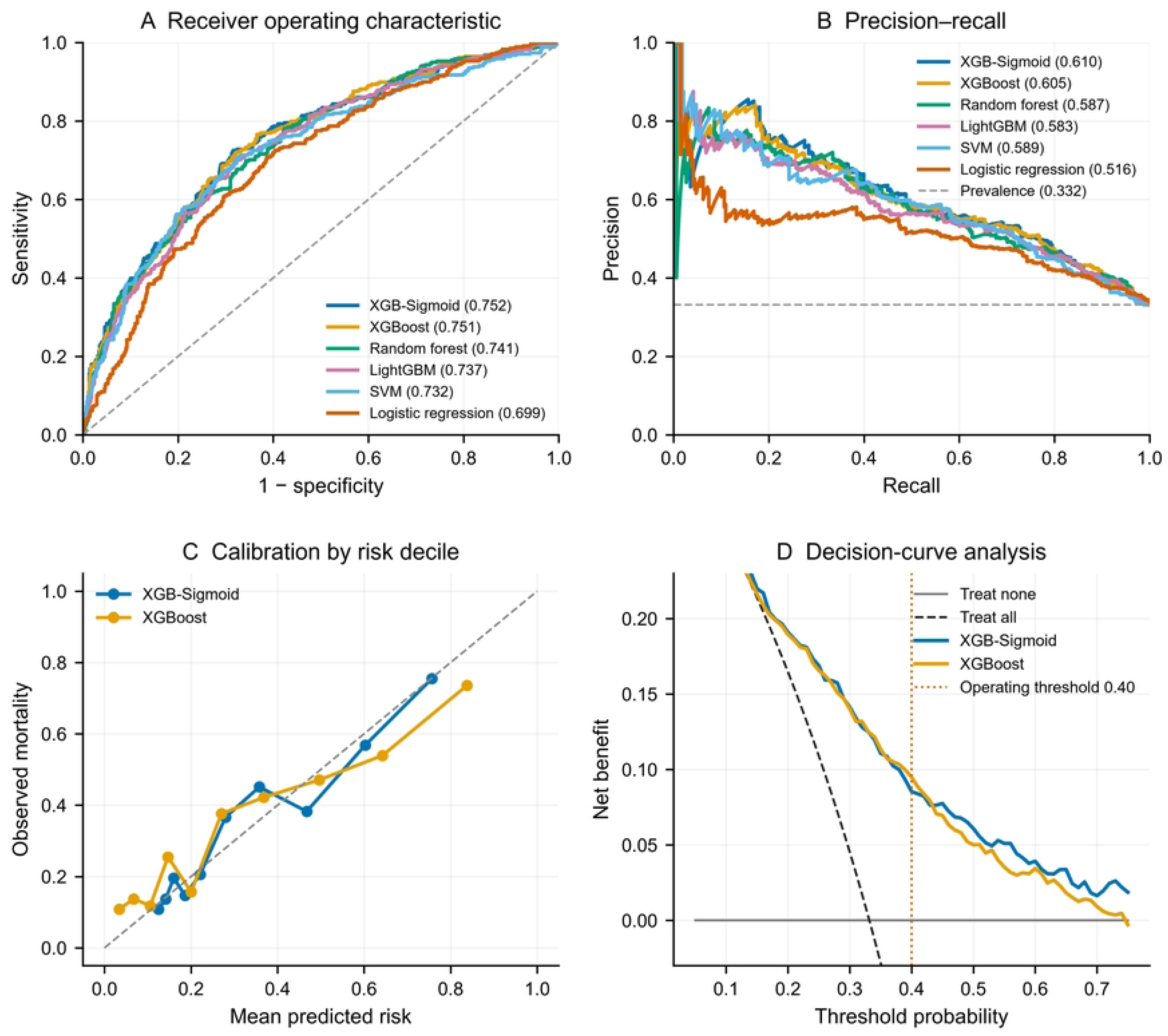
Model discrimination, calibration, and decision-curve analysis for internal validation of XGBoost. (A) Receiver operating characteristic curves, with ROC AUC values in the legend. (B) Precision–recall curves; the horizontal dashed line indicates the event proportion of 0.332. (C) Mean predicted risk and observed mortality by predicted-risk decile before and after calibration; the diagonal indicates perfect calibration. (D) Decision curves against treat-all and treat-none strategies. All panels use pooled out-of-fold predictions. Selected model curves are shown for clarity; Table 2 reports all models. ROC, receiver operating characteristic; AUC, area under the curve; PR, precision–recall; XGB-Sigmoid, sigmoid-calibrated XGBoost; SVM, support vector machine.

**Table 2.** Performance comparison of candidate models in pooled out-of-fold internal validation.

| Model | AUC [95%<br>CI] | AP | Brier | Calibration<br>intercept | Calibration<br>slope |
| --- | --- | --- | --- | --- | --- |
| XGB-Sigmoid | 0.752 [0.720,<br>0.784] | 0.610 | 0.182 | -0.011 | 0.966 |
| XGBoost | 0.751 [0.718,<br>0.781] | 0.605 | 0.186 | -0.127 | 0.670 |
| Random forest | 0.741 [0.709,<br>0.773] | 0.587 | 0.207 | -0.618 | 1.585 |
| LightGBM | 0.737 [0.704,<br>0.770] | 0.583 | 0.226 | -0.199 | 0.247 |
| Support vector<br>machine | 0.732 [0.699,<br>0.766] | 0.589 | 0.188 | -0.114 | 0.809 |
| Logistic<br>regression | 0.699 [0.665,<br>0.733] | 0.516 | 0.229 | 0.529 | 7.237 |
| BPNN | 0.682 [0.646,<br>0.717] | 0.544 | 0.274 | -0.409 | 0.101 |
| KNN | 0.706 [0.672,<br>0.739] | 0.546 | 0.204 | -0.401 | 0.103 |
| SAPS II | 0.651 [0.615,<br>0.685] | 0.467 | — | — | — |
| First SOFA | 0.615 [0.579,<br>0.652] | 0.436 | — | — | — |

**Table 3.** Exploratory internal operating characteristics and risk tertiles for the calibrated XGBoost model.

A. Exploratory operating characteristics
| Threshold | Sensitivity | Specificity | PPV | NPV | F1 | Net benefit |
| --- | --- | --- | --- | --- | --- | --- |
| 0.225 | 0.793 | 0.583 | 0.486 | 0.850 | 0.602 | 0.182 |
| 0.275 | 0.728 | 0.677 | 0.528 | 0.834 | 0.612 | 0.160 |
| 0.400 | 0.524 | 0.802 | 0.567 | 0.772 | 0.545 | 0.085 |

B. Exploratory risk tertiles
| Risk stratum | N | Deaths | Observed mortality | 95% CI |
| --- | --- | --- | --- | --- |
| Low | 340 | 48 | 14.1% | 10.8% to 18.2% |
| Intermediate | 339 | 99 | 29.2% | 24.6% to 34.3% |
| High | 340 | 191 | 56.2% | 50.9% to 61.4% |

### Exploratory comparison of static and static-plus-dynamic feature sets

The static-only arm included 44 candidate variables, whereas the static-plus-dynamic arm included 174. Their AUCs were 0.681 (95% CI 0.646–0.714) and 0.752 (0.724–0.787), respectively (Table 4). The paired AUC difference was 0.071 (95% CI 0.042–0.109). Average precision increased from 0.499 to 0.619, while the Brier score decreased from 0.202 to 0.181.

**Table 4.** Exploratory two-arm ablation comparison in the MIMIC-IV development cohort.

| Feature set | Candidate variables | AUC (95% CI) | Average precision / Brier |
| --- | --- | --- | --- |
| Static | 44 | 0.681 (0.646–0.714) | 0.499 / 0.202 |
| Static + dynamic | 174 | 0.752 (0.724–0.787) | 0.619 / 0.181 |

### Model interpretation

Urine-output measures, Charlson Comorbidity Index, base-excess slope, last systolic blood pressure, and last norepinephrine dose were prominent in the SHAP analysis (Fig 4). Features derived from the 72-hour course accounted for 85.2% of total mean absolute attribution. Level or summary features accounted for 53.2%, and trajectory or variability features for 32.0%.

**Fig 4.**
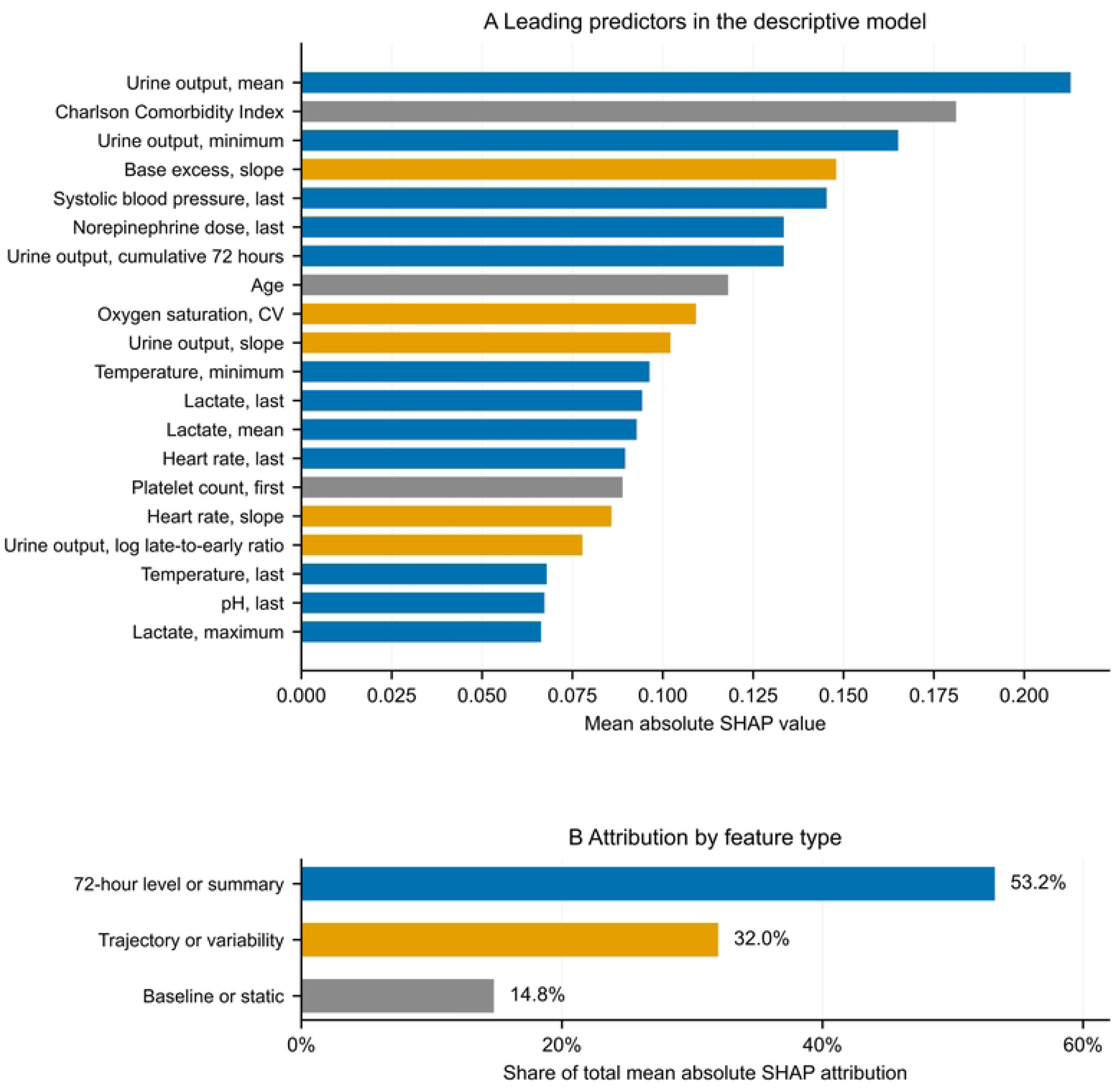
SHAP interpretation of the descriptive full-data XGBoost model. (A) The 20 predictors with the largest mean absolute SHAP values in the XGBoost model fitted to all 1019 development patients. (B) Attribution proportions across all 45 predictors, grouped by feature type. Blue denotes 72-hour level or summary metrics, orange denotes trajectory or variability metrics, and gray denotes baseline or static metrics. Attributions describe the fitted model rather than incremental predictive value. SHAP, Shapley additive explanations; CV, coefficient of variation. For the dynamic predictors, first and last refer to the first and last observed values within the 72-hour window; slope is expressed per 12-hour interval (S1 Text).

### External model performance

The locked model achieved an external AUC of 0.713 (95% CI 0.687–0.739), average precision of 0.536 (0.491–0.582), and Brier score of 0.185 (0.174–0.197; Fig 5). Mean predicted risk was 27.3%, compared with observed mortality of 29.5%. The calibration intercept was −0.109 and the slope was 0.744.

**Fig 5.**
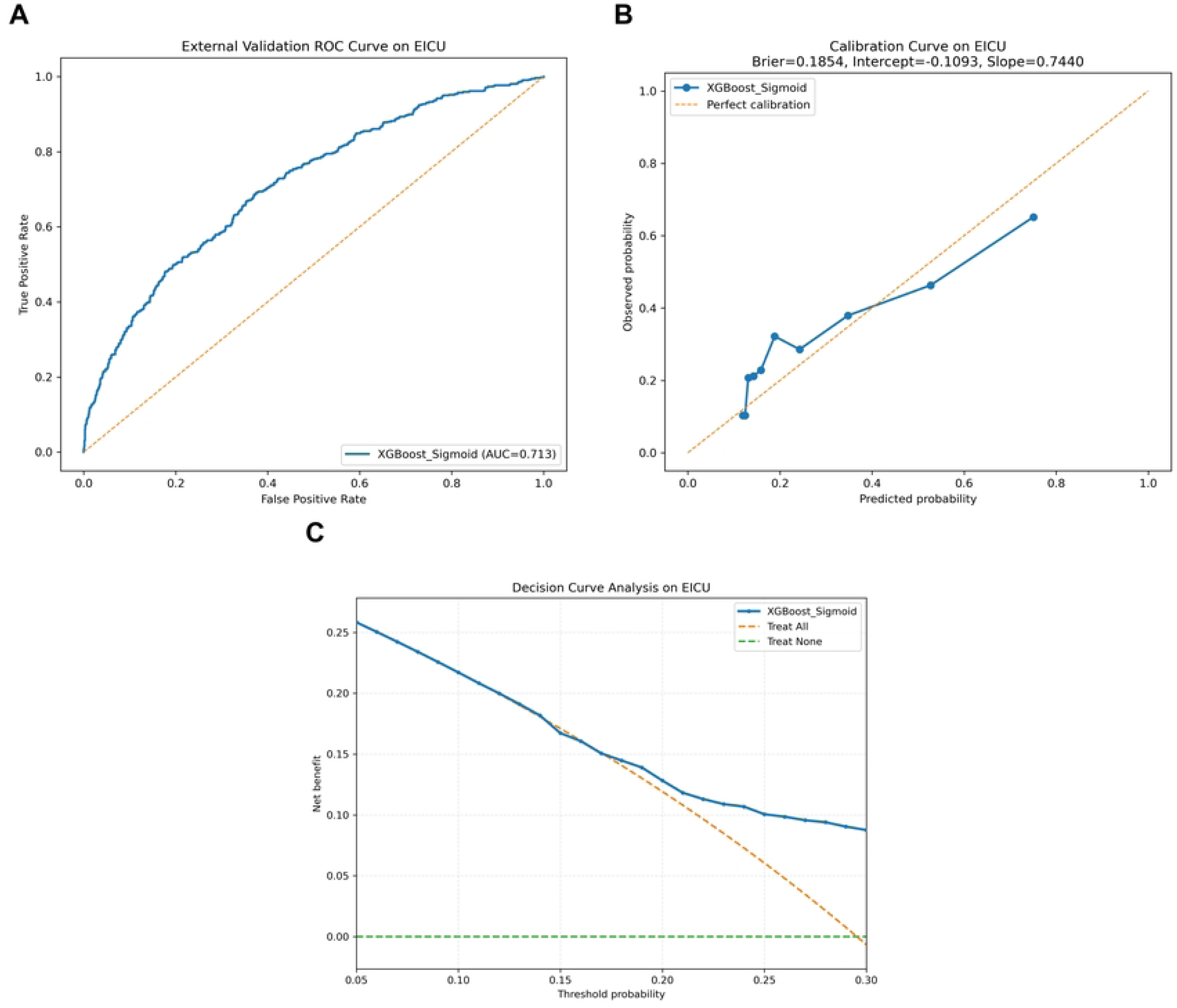
External validation performance of the locked model in eICU-CRD. (A) Receiver operating characteristic curve. (B) Mean predicted risk and observed mortality across risk groups; the diagonal indicates perfect calibration. (C) Decision-curve analysis over threshold probabilities of 0.05–0.30. Dashed reference curves represent treat-all and treat-none strategies. The model was applied to eICU-CRD without refitting or recalibration. ROC, receiver operating characteristic; AUC, area under the curve.

Across the evaluated 0.05–0.30 threshold-probability range, model net benefit exceeded treat-none but did not consistently exceed treat-all. Mortality in external low-, intermediate-, and high-risk tertiles was 13.7% (80/583), 27.5% (160/581), and 47.3% (276/583), respectively (Table 5). These strata were defined using the external predicted-risk distribution.

**Table 5.** Mortality in risk tertiles of the eICU-CRD external validation cohort, defined by predicted risk.

| Risk stratum | N | Deaths | Observed mortality |
| --- | --- | --- | --- |
| Low | 583 | 80 | 13.7% |
| Intermediate | 581 | 160 | 27.5% |
| High | 583 | 276 | 47.3% |

## Discussion

The first three days of intensive care provide an opportunity to reassess prognosis using the observed course of illness. In patients with sepsis and pre-existing hypertension who had received vasoactive agents, our model combined this information with static clinical characteristics and retained moderate discrimination in an independent database. The mortality gradient across external risk groups indicates that the model preserved information about relative prognosis beyond the development setting. The exploratory feature-set comparison further supports the value of a broader representation of information accumulated during the first 72 hours in this selected population. The clinical contribution is therefore a structured assessment of subsequent mortality risk among patients still in the ICU at 72 hours, a stage at which admission risk estimates can be reconsidered in light of the intervening course.

The focus on pre-existing hypertension is clinically motivated. A history of hypertension provides a chronic vascular context in which acute pressure changes, organ function, and the need for circulatory support should be interpreted together [6–8]. An arterial pressure value alone does not describe the support required to achieve it or the accompanying organ response. SEPSISPAM illustrates this distinction. A higher pressure target did not improve overall survival, although renal-replacement therapy was less frequent among patients with chronic hypertension in the higher-target group; newly diagnosed atrial fibrillation was more frequent overall [9]. These findings suggest that pressure, renal outcomes, and treatment burden need not move in parallel. They support considering several dimensions of the observed course when studying prognosis in hypertensive patients. Our findings address that prognostic question, without establishing a preferred blood-pressure target or a hypertension-specific mechanism.

The selected predictors give the model a clinically interpretable basis. Age and Charlson Comorbidity Index describe aspects of pre-existing vulnerability, while blood pressure, urine output, acid-base status, and vasopressor exposure characterize the acute course. Their combination may help distinguish patients with similar pressure measurements but different organ responses or support requirements. Urine output was particularly relevant through both measurement levels and an early-to-late ratio, consistent with previous evidence that its prognostic information depends on the amount and duration of reduced output [25]. However, the recorded course reflects both illness and care. Changes in urine output or drug exposure can accompany changes in severity and clinical decisions, and the model does not separate these influences. SHAP makes their contribution to the fitted predictions visible [22], but correlated features may share information and their attributions cannot identify independent biological effects or the benefit of changing treatment [26–27].

Repeated observation adds a temporal dimension to risk assessment. Patients with similar admission findings may subsequently improve, remain dependent on support, or develop persistent abnormalities. Prior studies of vital-sign dynamics and time-series prediction have similarly examined the prognostic information accumulated during intensive care [14–15]. Our use of six 12-hour windows summarizes that course at a single landmark, rather than providing a continuously updating prediction. The exploratory comparison used the same patient folds and repeated the modeling procedure in each feature arm, supporting the observed difference between the two feature sets. The observed difference should be interpreted as the added predictive information provided by the broader 72-hour feature representation, which incorporated measurement levels and summaries as well as temporal changes and variability. Because the static arm also included treatment and organ-dysfunction variables, this comparison does not isolate the independent contribution of trajectory features alone.

Independent validation showed both the promise and the limits of using this information across settings. Risk ordering persisted, but absolute probabilities were less well calibrated. The external slope below one was consistent with excessive dispersion of predicted risk, while the lower mean prediction indicated underestimation on average. The jointly estimated regularized intercept does not independently measure that average difference. Harmonized variables and units improve comparability, yet differences in case mix, measurement, and care may still affect performance; their individual contributions were not assessed. These results favor evaluating discrimination and calibration together rather than treating an AUC as sufficient evidence of transportability [28–31]. Local calibration assessment is needed before probabilities guide decisions. Similarly, the external tertiles demonstrate separation within that cohort, without establishing that fixed internal risk-group boundaries would transfer unchanged.

Decision-curve analysis provides a framework for considering the trade-off between missed events and unnecessary actions [23–24]. In the external cohort, model net benefit exceeded treat-none across the evaluated threshold-probability range but did not consistently exceed treat-all. These findings do not establish a specific clinical action threshold or demonstrate an improvement in care. The proposed use of the model as an aid to clinical reassessment therefore remains to be evaluated prospectively, with the downstream consequences of acting on predicted risk explicitly defined.

The contribution should also be judged in relation to the prediction time and the information available. Previous sepsis and ICU models differ in population, observation window, and outcome [13–15,32–38], which limits direct comparison of published AUCs. Our comparisons with SAPS II and first SOFA included the advantage of observing the subsequent course and therefore do not isolate an algorithmic benefit. Paired comparisons likewise did not establish clear discrimination superiority of sigmoid-calibrated XGBoost over uncalibrated XGBoost or random forest. The study instead brings together a defined clinical population, a fixed reassessment point, and independent evaluation of several aspects of prediction performance. Transparent reporting of these components is necessary for judging whether the findings are reproducible and applicable [31,39–42]. Comparisons with simpler approaches using the same prediction time and available information would help establish the practical value of this strategy.

Several limitations should be considered. First, this retrospective study was restricted to patients who remained in the ICU for at least 72 hours and received vasoactive agents, which limits generalizability to broader sepsis populations and may introduce selection related to missing data and routine clinical measurement practices. Second, the large candidate-feature set and variation in feature selection may have increased model complexity and instability; the selected predictors should therefore not be interpreted as a definitive set of prognostic factors. Finally, although the model was evaluated in an independent database, differences in case mix and care practices may affect calibration, and prospective evaluation with local calibration is required before clinical implementation.

## Conclusions

Clinical information accumulated during the first 72 hours supported subsequent mortality risk stratification in patients with sepsis and pre-existing hypertension who had received vasoactive agents. The model retained moderate discrimination in an independent cohort and may help structure risk reassessment after the initial ICU course. Its clinical use requires local calibration assessment and prospective evaluation of the decisions prompted by its predictions.

## Data Availability

No new patient-level data were generated by this study. The data analyzed in this study were obtained from two existing databases available through PhysioNet: MIMIC-IV version 3.1 (https://doi.org/10.13026/kpb9-mt58) and the eICU Collaborative Research Database version 2.0 (https://doi.org/10.13026/C2WM1R). Access to these databases requires completion of the applicable PhysioNet credentialing process, training, and data use agreements. The authors did not receive any special privileges in accessing these data, and other qualified researchers can obtain access under the same conditions through PhysioNet. Because the patient-level data are subject to PhysioNet data use agreements, the authors are not permitted to redistribute them. Aggregate results underlying the reported findings are provided in the manuscript and Supporting Information. Author-generated analysis code and reproduction instructions will be made publicly available upon publication.

## Supporting information

S1 Text. Definitions and calculations of dynamic predictors.

**S2 Fig. Pre-imputation missingness among the 30 candidate predictors with the highest missing proportions in the MIMIC-IV development cohort. The 20% exclusion criterion applied to each admission record, not to individual variables. The dashed 20% line is a visual reference for variable-level missingness, not a variable-exclusion threshold.**

S3 Table. Baseline and early clinical characteristics of the eICU-CRD external validation cohort.

